# Lineage H Rift Valley Fever Virus Drives Deadly 2025 Outbreak in Northern Senegal

**DOI:** 10.1101/2025.10.08.25337605

**Authors:** Moussa Moïse Diagne, Gamou Fall, Abiboulaye Sall, Bocar Sow, Ndeye Awa Ndiaye, Alioune Gaye, Mamadou Sarr Ndao, Aboubacry Gaye, El Hadji Ndiaye, Seynabou Mbaye Ba Souna Diop, Safiétou Sankhe, Mouhamed Kane, Seynabou Ndiaye, Ousmane Faye, Yoro Sall, Mamadou Aliou Barry, Ibou Gueye, Marie Henriette Dior Ndione, Boly Diop, Ousmane Cisse, Joseph R A Fitchett, Ibrahima Dia, Cheikh Loucoubar, Ndongo Dia, Mawlouth Diallo, Boubacar Diallo, Ibrahima Soce Fall, Mamadou Ndiaye, Diawo Diallo, Abdourahmane Sow, Oumar Faye

## Abstract

In late September 2025, a deadly Rift Valley fever outbreak in northern Senegal caused 44 cases and 8 deaths. Sequencing confirmed lineage H persistence with three novel polymerase mutations suggesting adaptation. Ongoing concurrent One Health investigations target vectors, livestock, and human exposure to identify risk factors and strengthen control.

## Introduction and Outbreak Context

Rift Valley fever virus (RVFV), a mosquito-borne phlebovirus of the family *Phenuiviridae*, was first described in Kenya in 1931 [1]. Since then, the virus has caused repeated epidemics in Africa and the Arabian Peninsula, including major outbreaks in Egypt (1977), East Africa (1997–98, 2006–07), Saudi Arabia and Yemen (2000) [2]. In West Africa, recurrent epidemics have been reported in Mauritania, Niger, Mali, and Senegal [3,4].

Senegal has a well-documented history of RVF outbreaks. The first major outbreak occurred in 1987, at the frontier of Senegal and Mauritania, producing approximately 1,500 human cases and more than 200 deaths [5]. Additional outbreaks or sporadic cases were reported in 1993–94, 1998–99, 2002–03, 2012, and 2013–14 but the impact was very minor without death [6]. In recent years, genomic studies have shown that lineage H, previously detected in Southern Africa, became established in Senegal from 2020 onward, replacing older West African lineages [7,8].

In September 2025, an outbreak of RVF was confirmed in the Saint-Louis region of northern Senegal. On 20 September, the first confirmed cases were identified at the regional hospital. By 26 September, eleven cases had been confirmed, including four deaths. As investigations expanded, the outbreak scale grew rapidly and 2 more regions (Matam and Louga were affected). The Ministry of Health classified the event as a severe epidemic requiring national-level coordination and activated the Regional Epidemic Management Committee along with an Incident Management System to oversee case management, contact tracing, and risk communication (Ministry of Health, Senegal, Situation Report No. 1, 26 Sept 2025). By 07 October, surveillance data indicated more than 600 suspected cases, of which 100 were confirmed by RT-qPCR and 19 by IgM serology, giving a total of 119 laboratory-confirmed infections. At that date, 15 deaths had been reported in total, corresponding to a high case fatality proportion among confirmed cases.

Given the severe clinical presentation, the Institut Pasteur de Dakar (IPD) conducted a rapid genomic sequencing of clinical isolates to assess whether viral genetic changes might account for the outbreak’s lethality.

## Methods

RNA from RT-qPCR–confirmed samples was sequenced on an Illumina iSeq100 using the Twist CVRP hybrid-capture workflow, following manufacturer’s instructions as previously described [8]. For genome assembly, raw FASTQ reads were trimmed with Trimmomatic (v0.39), and L, M, and S segments aligned to references using BWA MEM (v0.7.17). Consensus sequences were generated with iVar (v1.3.1) under a 50% allele frequency and ≥10× depth. BAM files were annotated with LoFreq indelqual, and SNVs/indels called with LoFreq (v2.1.5). For phylogenetic analyses, the 2025 outbreak genomes were combined with all complete and nearly complete RVFV genomes available in GenBank, aligned with MAFFT, and maximum-likelihood trees reconstructed with IQ-TREE using ModelFinder-selected models and 1,000 ultrafast bootstrap/SH-aLRT replicates. Trees were visualized in iTOL [9]. Mutations were identified from nucleotide alignments in AliView [10], highlighting lineage-specific changes. Codon display and amino acid translation distinguished synonymous from nonsynonymous substitutions, confirmed with MEGA using the Nei– Gojobori method and a codon-based Z-test of selection.

## Results and Discussion

Nine RVFV genomes spanning the L, M, and S segments were assembled and coverage exceeded 90% for five complete genomes (S1, S2, S3, S4, S8), while other samples showed partial segmental coverage (Table 1). For downstream analyses, we retained only high-quality sequences with minimum depth ≥ 15×, resulting in complete L/M/S genomes from S1, S2, S3, S4, and S8, and the M segment from S5.

**Table 1.** Sequencing coverage of RVFV genomes from the 2025 outbreak in Senegal.

| ID-Sequencing | ID-Diagnostic | RVFV Segment | Coverage (%) | Depth (x) |
| --- | --- | --- | --- | --- |
| S1 | 471872 | L | 99.86 | 1813.38 |
|  |  | M | 99.85 | 2645.57 |
|  |  | S | 99.76 | 4959.01 |
| S2 | 473088 | L | 99.86 | 2416.7 |
|  |  | M | 99.82 | 4182.69 |
|  |  | S | 99.7 | 4231.65 |
| S3 | 471871 | L | 99.81 | 64.786 |
|  |  | M | 99.41 | 91.3745 |
|  |  | S | 99.41 | 81.198 |
| S4 | 473086 | L | 99.81 | 850.33 |
|  |  | M | 99.67 | 902.365 |
|  |  | S | 98.93 | 2006.94 |
| S5 | 473084 | L | 69.94 | 9.18785 |
|  |  | M | 95.73 | 12.2494 |
|  |  | S | 94.38 | 9.2145 |
| S6 | 473113 | L | 0.0 | 0.0 |
|  |  | M | 0.0 | 0.0388674 |
|  |  | S | 0.0 | 0.0 |
| S7 | 471869 | L | 31.48 | 4.11438 |
|  |  | M | 60.95 | 6.71995 |
|  |  | S | 44.79 | 5.27395 |
| S8 | 473112 | L | 99.81 | 100.237 |
|  |  | M | 99.43 | 173.675 |
|  |  | S | 99.41 | 131.082 |
| S9 | 471625 | L | 16.94 | 2.84971 |
|  |  | M | 46.0 | 5.1861 |
|  |  | S | 57.51 | 7.4911 |
| S10 | 471624 | L | 51.76 | 7.1008 |
|  |  | M | 94.95 | 13.1504 |
|  |  | S | 78.05 | 8.09735 |

Comparative analysis showed that the 2025 outbreak genomes were highly conserved and shared more than 99% nucleotide identity with previously sequenced Senegalese isolates from Fatick (2020) and Matam (2022) [7,8]. Phylogenetic reconstruction confirmed that the outbreak strains clustered within the West African branch of lineage H, demonstrating continued local circulation rather than the introduction of a novel genotype (Figure 1).

**Figure 1 legend.**
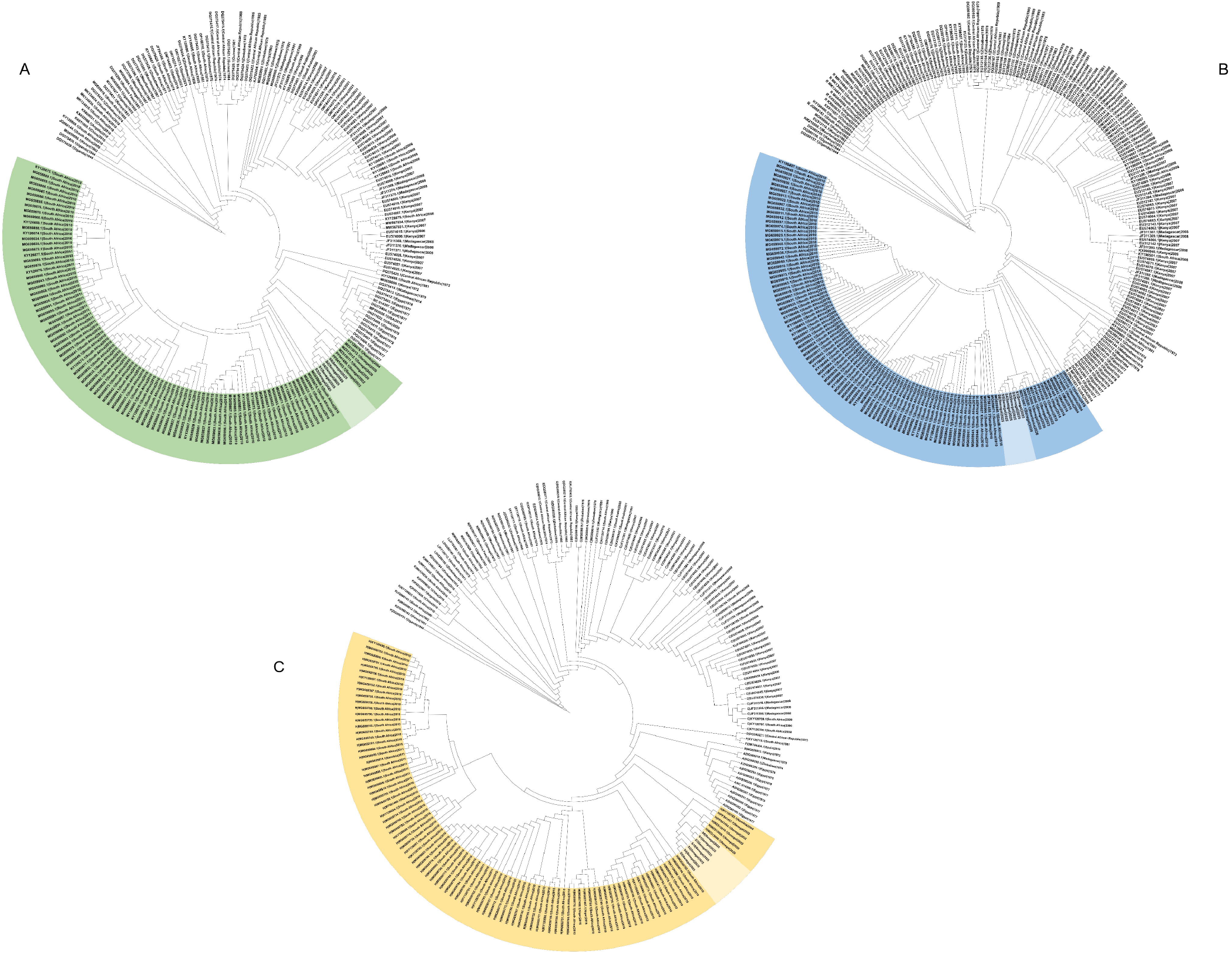
Maximum-likelihood phylogenetic trees of the L (A), S (B) and M (C) segments constructed from all complete and nearly complete Rift Valley fever virus genomes available in GenBank. Sequences belonging to lineage H (highlighted) include the 2025 Senegal outbreak genomes, which cluster with earlier isolates from Fatick (2020), Matam (2022) and Mauritania (2020), indicating ongoing local circulation rather than a new introduction.

Selection pressure analysis showed different evolutionary patterns across the genomes. When compared with the reference strain ZH-548 M12, the outbreak sequences showed clear signs of positive selection (Z ≈ –3.4 to –4; p < 0.01), meaning they accumulated more amino acid changes than expected. In contrast, most of the recent outbreak isolates were under purifying selection (Z > 0), indicating that their proteins remain highly conserved. Looking at changes over time, comparisons between 2020, 2022, and 2025 genomes suggested moderate adaptive trends, with slightly negative Z values (–1.5 to –1.8) but non-significant p-values (0.07–0.13). Full results are provided in the Supplementary Tables S1-S3.

Most amino acid substitutions were conservative, including R137K in NSs, K1111R in NSm, and multiple changes in the L polymerase (R158K, R493K, R1926K, E177D, V1760I) (Table 2). These substitutions involve residues of similar biochemical properties and are unlikely to significantly alter protein function [11].

**Table 2.** Amino Acid Mutations Identified in RVFV Lineage H Strains from the 2025 Senegal Outbreak.

| Gene / Segment | Mutation (AA change) | Type (Conservative vs Non-conservative) | Likely Effect on Virus | Conclusion |
| --- | --- | --- | --- | --- |
| NSs (S segment) | R137K (Arg → Lys) | Conservative (both basic) | Minimal effect on interferon antagonism; still functional | Marker mutation, not major functional impact |
| NSm (M segment) | K1111R (Lys → Arg) | Conservative (both basic) | Anti-apoptotic function likely preserved | Marker mutation, little phenotypic change |
| L polymerase | D11N (Asp → Asn) | Non-conservative (charge lost) | Possible subtle change in N-terminal endonuclease (cap-snatching) | May fine-tune replication efficiency |
| L polymerase | M120T (Met → Thr) | Non-conservative (hydrophobic → polar) | Could alter local folding near endonuclease | Potential minor replication effect |
| L polymerase | R158K, R493K, R1926K (Arg → Lys) | Conservative | Likely neutral for polymerase activity | Neutral substitutions |
| L polymerase | E177D (Glu → Asp) | Conservative | No significant effect (similar negative charge) | Neutral substitution |
| L polymerase | V1760I (Val → Ile) | Conservative | No major impact on hydrophobic core | Neutral substitution |
| L polymerase | Y1852H (Tyr → His) | Non-conservative (aromatic → basic) | Could affect C-terminal interactions or stability | Worth monitoring; possible subtle effect |
| Lineage H (general) | Shared synonymous mutations | Neutral | Useful for phylogenetic tracking | Serve as lineage markers |

Three non-conservative substitutions were identified in the L polymerase. The D11N mutation, located in the endonuclease domain, may subtly affect cap-snatching activity. The M120T substitution introduces a polar residue in place of a hydrophobic methionine in the N-terminal region, potentially influencing local folding or stability [12]. The Y1852H change replaces an aromatic tyrosine with a basic histidine in the C-terminal region, which could affect ribonucleoprotein interactions, replication fidelity, and adaptability [13]. The NSs protein, the main interferon antagonist [14], carried only one conservative substitution, suggesting preserved function. NSm also retained conservative variation. Importantly, no amino acid changes were observed in the glycoproteins Gn and Gc at known neutralizing epitopes, suggesting that immune recognition and vaccine efficacy should remain unaffected [15].

Although most 2025 outbreak genomes showed purifying selection, the presence of moderate adaptive trends over time suggests that lineage H continues to evolve while maintaining strong functional conservation. This pattern likely contributes to its persistence in Senegal and underlines the need for ongoing genomic surveillance to track gradual adaptive changes.

Lineage H had already been flagged as having phenotypic traits associated with higher virulence. Indeed, lineage H caused large-scale epidemics in South Africa, Namibia, and Mauritania [6,8]. During the 2020 Mauritania outbreak, which resulted in 78 confirmed human cases and 25 deaths, partial NSs sequences (Genbank accession: ON052829) clustered within lineage H (Figure 1A) and showed 100 % nucleotide identity with the Senegal 2020 (Fatick) isolates, confirming regional persistence of this lethal strain. In addition, lineage H showed higher viral replication efficiency *in vitro* compared with other epidemic lineages (G and C) and carried non-conservative mutations of possible functional significance [8]. The severe cases and fatalities now observed in Senegal are consistent with this earlier pattern and underscore the need for continued genomic and functional surveillance of lineage H viruses.

Taken together, the 2025 outbreak confirms the persistence of lineage H in West Africa, with highly conserved genomes punctuated by a few non-conservative polymerase substitutions that may subtly influence viral fitness. While immune recognition remains preserved, the occurrence of severe cases and fatalities underscores the need for sustained genomic and functional surveillance. These findings also reinforce the importance of integrated genomic and epidemiological monitoring to detect shifts in viral replication dynamics before they translate into larger outbreaks. The ongoing One Health outbreak investigations in northern Senegal aim to shed light on key ecological drivers, mosquito abundance and infection rates, livestock amplification, and human exposure patterns, to identify risk factors and inform strategies for better outbreak control.

## Data Availability

Consensus genomes (2025 Senegal) submitted to GenBank [accessions pending].
Raw FASTQ files and analysis scripts available on request to Institut Pasteur de Dakar.

https://virological.org/t/molecular-characterization-of-rift-valley-fever-virus-from-the-2025-outbreak-in-northern-senegal/1006

## Supplementary

Supplementary Table S1. Pairwise selection pressure analyses of RVFV S segment genomes Supplementary Table S2. Pairwise selection pressure analyses of RVFV L segment genomes Supplementary Table S3. Pairwise selection pressure analyses of RVFV M segment genomes

## Acknowledgments

We thank the healthcare workers from the Senegalese Ministry of Health for their dedication to outbreak management. We are also grateful to the teams from the different departments at Institut Pasteur de Dakar.

## Data availability

Consensus genomes (2025 Senegal) submitted to GenBank [accessions pending]. Raw FASTQ files and analysis scripts available on request to Institut Pasteur de Dakar.

